# Covid-19 Vaccine Effectiveness in Healthcare Personnel in six Israeli Hospitals (CoVEHPI)

**DOI:** 10.1101/2021.08.30.21262465

**Authors:** Mark A. Katz, Efrat Bron Harlev, Bibiana Chazan, Michal Chowers, David Greenberg, Alon Peretz, Sagi Tshori, Joseph Levy, Mili Yacobi, Avital Hirsch, Doron Amichay, Ronit Weinberger, Anat Ben Dor, Elena Keren Taraday, Dana Reznik, Chen Barazani Chayat, Dana Sagas, Haim Ben Zvi, Rita Berdinstein, Gloria Rashid, Yonat Shemer Avni, Michal Mandelboim, Neta Zuckerman, Nir Rainy, Amichay Akriv, Noa Dagan, Eldad Kepten, Noam Barda, Ran D. Balicer

**Affiliations:** Clalit Research Institute, Innovation Division, Clalit Health Services, Ramat Gan, Israel; School of Public Health, Faculty of Health Sciences, Ben Gurion University of the Negev, Beer Sheva, Israel; University of Michigan School of Public Health, Ann Arbor, MI, USA; Schneider Children’s Medical Center of Israel, Petach Tikvah, Israel; Infectious Diseases and Infection Control Unit, Ha’Emek Medical Center, Afula, Israel; Rappaport Faculty of Medicine, Technion. Haifa, Israel; Infectious Diseases, Meir Medical Center, Kfar Saba, Israel; Sackler School of Medicine, Tel-Aviv University, Tel Aviv, Israel; Pediatric Infectious Disease Unit the Pediatric Division, Soroka University Medical Center, Beer Sheva, Israel; Faculty of Health Sciences, Ben Gurion University of the Negev, Beer Sheva, Israel; Occupational Medicine Clinic, Rabin Medical Center, Petah Tikva, Israel; Research Authority, Kaplan Medical Center, Rehovot, Israel; The Faculty of Medicine, Hebrew University of Jerusalem, Jerusalem, Israel; Clalit Central Laboratory, Clalit Health Services, Tel Aviv, Israel; Multidisciplinary laboratory, Schneider Children’s Medical Center of Israel, Petach Tikvah, Israel; Clinical Microbiology Laboratory, Ha’Emek Medical Center, Afula, Israel; Microbiology Department, Rabin Medical Center, Petah Tikva, Israel; Microbiology Department, Kaplan Medical Center, Rehovot, Israel; Department of Clinical Laboratories, Meir Medical Center, Kfar Saba, Israel; Virology, Soroka University Medical Center, Beer Sheva, Israel; Central Virology Laboratory, Chaim Sheba Medical Center, Ministry of Health, Ramat Gan, Israel; Department of Epidemiology and Preventive Medicine, Sackler Faculty of Medicine, School of Public Health, Tel Aviv University, Tel Aviv, Israel; Laboratory Division, Shamir Medical Center, Zerifin, Israel; Software and Information Systems Engineering, Ben Gurion University, Beer Sheva, Israel; Department of Biomedical Informatics, Harvard Medical School, Boston, MA, USA; Ivan and Francesca Berkowitz Family Living Laboratory at Harvard Medical School, Boston, MA, USA

## Abstract

**Background:** Methodologically rigorous studies on Covid-19 vaccine effectiveness (VE) in preventing SARS-CoV-2 infection are critically needed to inform national and global policy on Covid-19 vaccine use. In Israel, healthcare personnel (HCP) were initially prioritized for Covid-19 vaccination, creating an ideal setting to evaluate real-world VE in a closely monitored population.

**Methods:** We conducted a prospective study among HCP in 6 hospitals to estimate the effectiveness of the BNT162b2 mRNA Covid-19 vaccine in preventing SARS-CoV-2 infection. Participants filled out weekly symptom questionnaires, provided weekly nasal specimens, and three serology samples – at enrollment, 30 days and 90 days. We estimated VE against PCR-confirmed SARS-CoV-2 infection using the Cox Proportional Hazards model and against a combined PCR/serology endpoint using Fisher’s exact test.

**Findings:** Of the 1,567 HCP enrolled between December 27, 2020 and February 15, 2021, 1,250 previously uninfected participants were included in the primary analysis; 998 (79.8%) were vaccinated with their first dose prior to or at enrollment, all with Pfizer BNT162b2 mRNA vaccine. There were four PCR-positive events among vaccinated participants, and nine among unvaccinated participants. Adjusted two-dose VE against any PCR-confirmed infection was 94.5% (95% CI: 82.6%-98.2%); adjusted two-dose VE against a combined endpoint of PCR and seroconversion for a 60-day follow-up period was 94.5% (95% CI: 63.0%-99.0%). Five PCR-positive samples from study participants were sequenced; all were alpha variant.

**Interpretation:** Our prospective VE study of HCP in Israel with rigorous weekly surveillance found very high VE for two doses of Pfizer BNT162b2 mRNA vaccine against SARS-CoV-2 during a period of predominant alpha variant circulation.

**Funding:** Clalit Health Services

## Introduction

Mass vaccination is considered the most important strategy to achieve sustained mitigation of the threat posed by Covid-19, by preventing morbidity and mortality and reducing SARS CoV-2 transmission.^1^ While clinical trials^2^ and real-world effectiveness studies^3^ have demonstrated the effectiveness of Covid-19 vaccines in preventing SARS-CoV-2 infections, most of these studies were not designed to accurately assess the extent by which vaccines reduce asymptomatic infection, a modality that has been shown to play an important role in virus transmission.^4^

Evidence is building that Covid-19 vaccines reduce asymptomatic infection and onward viral transmission. Two prospective VE studies among healthcare personnel (HCP), from the UK^5^ and the US,^6^ both of which conducted routine sampling of participants, found high two-dose VE (85% and 91%, respectively) against any infection with the Pfizer BNT162b2 mRNA vaccine. While these studies were methodically rigorous and robust in sample size, the UK study^5^ collected PCR samples from asymptomatic participants biweekly (although frontline workers were tested by lateral flow device twice a week) and therefore may have missed asymptomatic infections, which may cause viral shedding for only a few days^7^. In addition, neither study used serology testing to detect new infections that could have been missed by respiratory swabbing. Studies from Israel and the US have shown that viral RNA load is lower in infected vaccinated persons compared to unvaccinated persons, suggesting a lower risk of transmission in vaccinated individuals.^6,8^ Studies from Scotland and England have demonstrated a reduction in secondary infections among families of vaccinated persons compared to families of unvaccinated individuals.^9,10^

In Israel, a the national Covid-19 vaccination campaign began on December 20, 2020, in which HCP were part of the first group to be prioritized for vaccination; all HCP in the country were offered two doses of the Pfizer BNT162b2 vaccine separated by 21 days. We aimed to evaluate VE against any SARS-CoV-2 infection by conducting a prospective cohort study among HCP in six hospitals in Israel.

## Methods

### Setting

Clalit Health Services (CHS) is the largest of four integrated payer/provider health care organizations in Israel, with 4.7 million members (52% of the population). CHS operates 14 hospitals in Israel. For this study we enrolled HCP from six CHS hospitals: four in central Israel (Rabin Medical Center, Schneider Children’s Medical Center of Israel, Meir Medical Center and Kaplan Medical Center), one in northern Israel (Ha’Emek Medical Center) and one in southern Israel (Soroka University Medical Center). All hospitals are managed by CHS and mostly staffed by HCP insured by CHS. CHS has maintained fully digitalized electronic medical records (EMR) for over 20 years. which contains comprehensive data on all aspects of medical care. Data related to all COVID- 19 PCR tests and vaccine administration in Israel is collected centrally by the Israeli Ministry of Health (MoH) and is updated daily into CHS’s EMR system.

### Study Design

We conducted a prospective cohort study, designed to evaluate the effectiveness of the Pfizer BNT162b2 mRNA Covid-19 vaccine in preventing SARS-CoV-2 infections among HCP. The design was based in part on a World Health Organization (WHO)/Europe VE guidance document.^11^ To optimize the chances of detecting asymptomatic infections and identifying all symptomatic infections, we collected weekly nasal swabs from all participants, administered weekly symptom questionnaires, and collected serology samples at three different points during the follow-up period.

### Population

We offered enrollment to all HCP who were insured by CHS, working at the participating hospitals, and were eligible to receive the Covid-19 vaccine according to the Ministry of Health (MoH) guidelines.^12^ At the time of enrollment, the Israeli MoH policy of delaying vaccine in previously infected individuals had not yet been clearly articulated, so previously infected HCP were offered enrollment in the study. Due to concerns that enrollment antibody testing would not be able to distinguish between previous infection and vaccination, we did not enroll HCP who had received their first dose of the vaccine more than 21 days prior to the enrollment date.

### Recruitment

We used CHS’s EMR to generate a list of all HCP who worked at any of the six study hospitals and were also insured by CHS. Recruitment started on December 27, 2020 and continued through February 15, 2021. Participants were recruited regardless of their intention to get vaccinated.

### Data Collection

At enrollment all participants had a serology specimen and a respiratory specimen collected, and completed a brief questionnaire about demographic information; occupational, community, and household exposures; attitudes about the Covid-19 vaccine; and symptoms in the past week. Every week, for 12 weeks following enrollment, participants were asked to complete an electronic symptom questionnaire with questions about whether they had any of 16 symptoms suggestive of suspected Covid-19 illness. In addition, participants provided weekly nasal or combined nasal and throat specimens, regardless of whether they had symptoms. Participants were instructed to seek medical care when symptomatic; SARS-CoV-2 testing within the Israeli healthcare system was conducted according to Israel MoH guidelines.^13^

Study staff were instructed to contact all participants who tested positive for SARS-CoV- 2 test after the positive test results became available, regardless of the reason for the test, and administer a brief questionnaire about symptoms the participant had experienced five days before and five days after the test was conducted.

Participants filled out three questionnaires (enrollment, weekly, follow-up of PCR-positive participants), which included questions about the following symptoms (comprised mainly of symptoms from the WHO case definition for suspected and probable Covid-19^14^): fever; a new or worsening cough; new or worsening shortness of breath; chills; new or worsening muscle aches; new loss of taste; new loss of smell; sore throat; vomiting; diarrhea; nausea; fatigue; headache; nasal congestion or runny nose; change in mental state.

Participants provided two additional blood samples for serology – one at 30 days and another at 90 days after enrollment (for participants unvaccinated at enrollment) or 30 and 90 days after the receipt of the first vaccine dose (for participants vaccinated prior to or at enrollment). In order to accommodate for varying HCP work schedules, study staff were instructed to collect serology samples within one week of the target date for the 30- and 90-day serology draws.

We extracted data on participants’ sex, age, population sector, socioeconomic status (SES), history of influenza vaccination in the past five years, and Covid-19 vaccination history from the CHS EMR, as previously described.^15^ We also extracted data on chronic medical conditions that were identified by the Centers for Disease Control and Prevention (CDC) as risk factors for severe Covid-19^16^, using International Classification of Diseases, Ninth Revision (ICD-9) codes from inpatient and outpatient records and internal patient registries.^15^ We extracted EMR data on SARS-CoV-2 PCR tests conducted before and during the study. Finally, to evaluate disease severity, we extracted data on hospitalizations among participants who tested positive for SARS-CoV-2 during the study period, for the one-month period following the positive test.

### Data Management

Data collection and management for the study were conducted using REDCap, a browser-based software system (Vanderbilt University, Nashville, TN, USA). Questionnaires were designed to be self-administered electronically through the internet, using computers or mobile telephones.

### Laboratory Testing

#### RT-PCR Testing

Respiratory specimens were tested by RT-PCR by the respective laboratories in five of the six hospitals, and by the CHS central laboratory (Supplementary Table 1).

#### Genomic Sequencing

PCR-positive samples from participants underwent genetic sequencing at the Israel National Virology Laboratory (NVL) and Shamir Medical Center (SMC). (Supplemental Methods 1)

#### Serology

Enrollment serology samples underwent testing at either one of the study hospitals or the CHS Central Laboratory for antibodies to SARS-CoV-2 using a combination of the Abbott SARS-CoV-2 nucleocapsid IgG test (Abbott Laboratories, Sligo, Ireland), the Liaison SARS-CoV-2 spike protein S1/S2 IgG test (DiaSorin, Centralino, Italy), and the Abbott SARS-CoV-2 IgG II Quant test (Abbott Laboratories, Sligo, Ireland). Serology specimens collected at 30 days and 90 days were tested with the Abbott SARS-CoV-2 nucleocapsid IgG test and either the Abbott SARS-CoV-2 IgG II Quant test or the Liaison SARS-CoV-2 spike protein S1/S2 IgG test. Because the latter two anti-spike protein antibody tests were likely to be positive for vaccinated participants, we only used results from the Abbott SARS-CoV-2 nucleocapsid IgG test to determine new infections. Supplemental Methods 2 outlines the algorithm used to determine serology results.

### Sample Size Considerations

Samples size considerations are described in Supplemental Methods 3.

### Assessment of vaccine effectiveness

As a primary outcome, we evaluated VE in preventing any PCR-confirmed Covid-19 among participants who were at least sevenJdays after receipt of the second vaccine dose compared to participants who had never been vaccinated. As secondary analyses, we estimated VE in preventing asymptomatic and symptomatic PCR-confirmed Covid- 19 separately, and two-dose VE 14 days after receipt of the second vaccine dose.. We defined asymptomatic infection as one in which the participant was PCR-positive and denied symptoms in the seven days before and five days after specimen collection.

Finally, we estimated VE in preventing any infection as defined by a combined outcome of either PCR-confirmed infection and/or seroconversion. For this analysis, participants with non-negative enrollment serology or non-negative 30-day serology results (Supplemental Methods 2), those who were vaccinated after enrollment, and those who received only one dose of vaccine, were excluded from the analysis. All participants were followed for 60 days - from day 30 to day 90 following their first vaccination (for participants vaccinated prior to or at enrollment) or from day 30 to day 90 following the day of enrollment (for unvaccinated participants). Participants who had a negative 30- day serology and a positive 90-day serology were considered to have seroconverted to SARS-CoV-2 during the follow-up period.

For our primary analyses, in order to ensure that we were evaluating VE in an infection-naïve population, we only included participants who were seronegative at enrollment and did not have PCR-confirmed SARS-CoV-2 infection at or prior to enrollment. We also excluded fully vaccinated participants who had PCR-confirmed SARS-CoV-2 infection prior to seven days after their second vaccine dose. (Figure 1). For our secondary analyses, we included less restrictive cohorts (Supplementary Methods 4).

**Figure 1.**
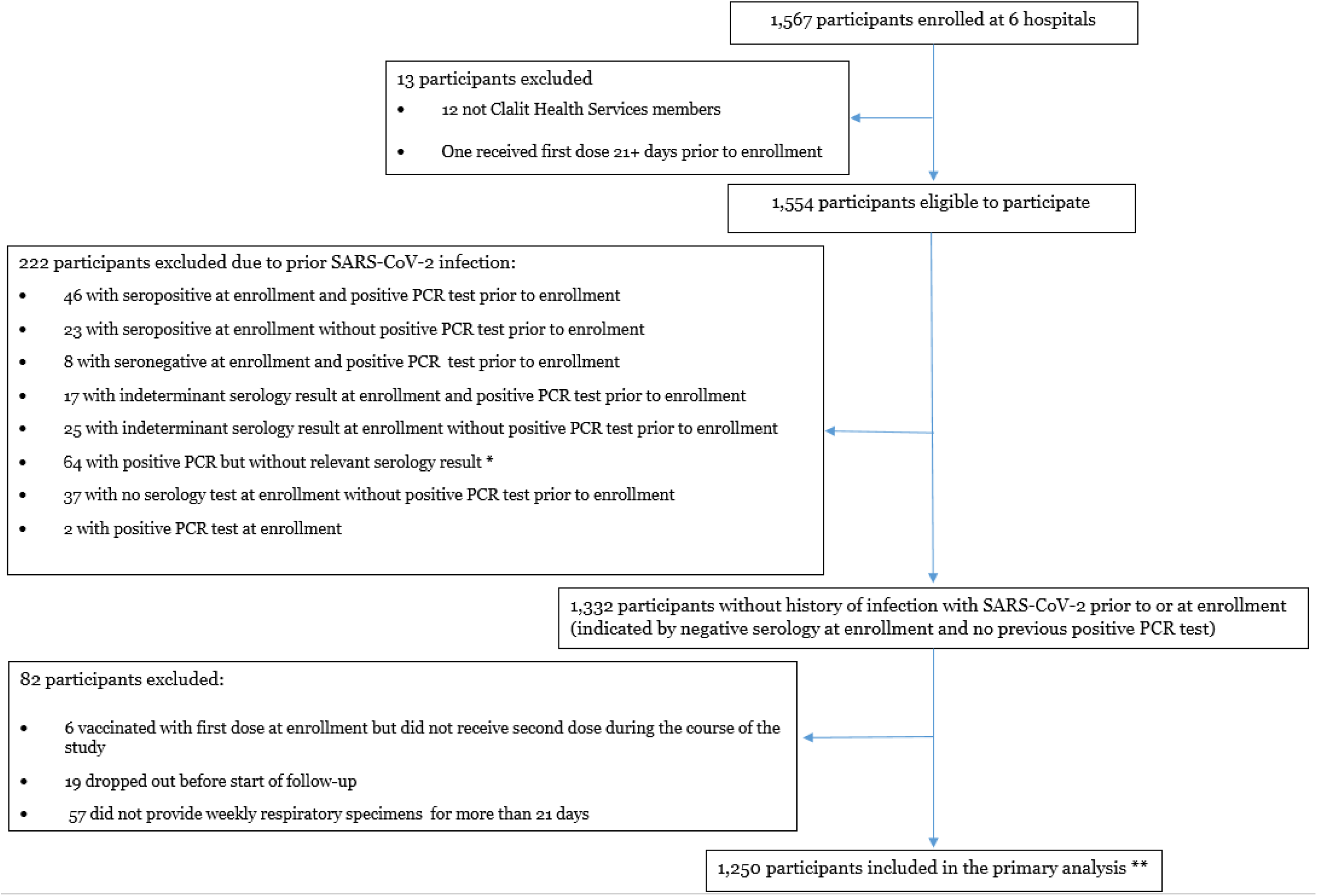
Study population and persons excluded to establish cohort for primary analysis, CoVEHPI. Notes: *If participants were already vaccinated before or at enrollment, and their enrollment serology was tested for anti-spike protein antibodies but not anti-nucleocapsid antibodies, their serology test was considered irrelevant (See Supplementary Methods 2) ** 9 PCR infections occurred among participants who were not eligible for the primary analysis due to non-negative serology or because the infections occurred among vaccinated participants before 7 days after they received their second vaccine dose

### Determination of follow-up time

We calculated follow-up time in person-days (PDs) from the day of enrollment. Participants were followed-up for 90 days from enrollment. For participants who were unvaccinated at enrollment, the time to infection was measured from the day of enrollment. For subjects who were enrolled as unvaccinated and chose to get vaccinated during the study, follow-up time was right-censored on the day the first vaccine dose was received. For subjects vaccinated before or at enrollment, the follow-up period started from the seventh day after they received the second vaccine dose. Participants infected during the study were followed until the date of their first PCR-positive test.

### Statistical analysis

The primary analysis was based on a Cox proportional hazards model, adjusted for the following covariates: age, sex, SES, population sector (Arab/Jewish) and occupation (physician/nurse or administrative/support staff). Hospital of employment was included as a random effect. In all analyses, two methods were used to account for fluctuations in the weekly Covid-19 infection rates in Israel.^17^ First, calendar time was used as the time scale of the Cox model. Second, as a sensitivity analysis, time from start of follow-up was used as the time scale of the Cox model, and a time-varying covariate with the weekly incidence of new COVID-19 cases in Israel was added to the model. VE was defined as one minus the hazard ratio between the vaccinated and unvaccinated. For the combined outcome of a positive PCR test or seroconversion, we applied Fisher’s exact test and VE was defined as one minus the odds ratio. For the additional analyses we employed the same modeling that we used for the primary analysis. There were no missing data related to the analyses.

### Ethics

The study protocol was reviewed and approved by the CHS Central Institutional Review Board All participants completed written informed consent in Hebrew. Small gifts (total value < $10 per participant) were given to participants at study milestones. Participants had access to results of all laboratory tests performed during the study.

## Results

We enrolled 1,567 HCP from the six hospitals between December 27, 2020 and February 15, 2021 (Supplementary Table 2). Of the 1567 enrolled participants, 1250 (79.8%) were included in the primary analysis (Figure 1). Overall, 222 (14.2%) participants were excluded in the primary analysis because they had a positive or indeterminate enrollment serology result, did not have relevant enrollment serology results; or had a PCR-positive test before or at enrollment.

Of the 1,250 participants in the primary analysis, 998 (79.8%) were vaccinated before or at enrollment. These participants received their second vaccine dose at a median of 22 days (interquartile range [IQR]: 22-22) after the first dose. Figure 2 shows the weeks participants received their first and second vaccine doses in the context of the national pandemic activity. Of the 252 participants who were unvaccinated at the time of enrollment, 201 (79.8%) received a first vaccine dose during the three-month follow-up period and thus did not complete the full follow-up period. The median time from enrollment to vaccination among participants who entered the study unvaccinated was 34 days (IQR: 16-51).

**Figure 2.**
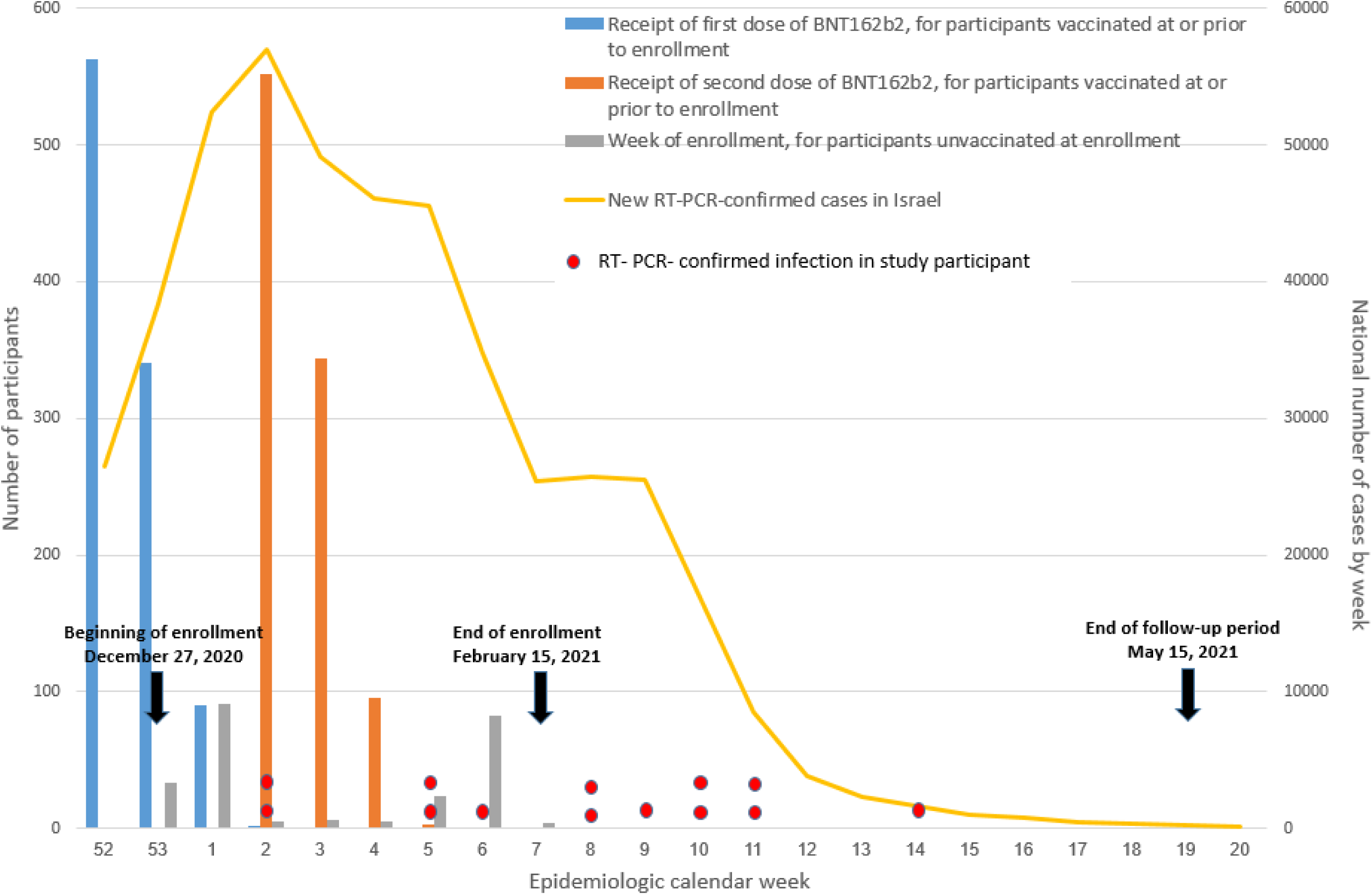
Week of first and second vaccination for vaccinated participants, week of enrollment for unvaccinated participants, RT-PCR-confirmed cases in study participants, and new weekly SARS-CoV-2 cases in Israel, December 2020-May 2021

Most vaccinated (77.3%) and unvaccinated (90.5%) participants were female. Vaccinated participants who were included in the primary analysis had a median age of 47 years (IQR: 38-57) compared to 37 (IQR: 31-47) in unvaccinated participants (Table 1). Less vaccinated participants were Arab compared to unvaccinated participants (5.7% vs. 11.5%, p<0.0001), and less vaccinated participants had low SES compared to unvaccinated participants (9.4% vs. 17.1%, P<0.0001). There were more physicians among vaccinated participants than unvaccinated participants (21.9% vs. 6.0%, p<0.0001). The percentages of participants who had contact with Covid-19 patients, had direct contact with patients and had at least one comorbidity were similar between vaccinated and unvaccinated patients, but more vaccinated patients received influenza vaccines in previous years compared to unvaccinated participants.

**Table 1.** Demographic and clinical characteristics of participants included in primary analysis, Covid-19 vaccine effectiveness in healthcare personnel in six Clalit Health Services hospitals in Israel (CoVEHPI), December 2020-May 2021

| Characteristic | All Participants<br>no. (%) | Vaccinated<br>no. (%) | Not<br>Vaccinated<br>no. (%) | P value |
| --- | --- | --- | --- | --- |
| Total | 1250 | 998 (79.8%) | 252 (20.2%) |  |
| Sex |  |  |  |  |
| Female | 999 (79.9%) | 771 (77.3%) | 228 (90.5%) | <0.0001 |
| Male | 251 (20.1%) | 227 (22.7%) | 24 (9.5%) |  |
| Age, Median (IQR) | 45 (36-55) | 47 (38-57) | 37 (31-47) |  |
| Age group |  |  |  |  |
| 18-34 | 266 (21.3%) | 167 (16.7%) | 99 (39.3%) | <0.0001 |
| 35-49 | 497 (39.8%) | 396 (39.7%) | 101 (40.1%) |  |
| 50-64 | 420 (33.6%) | 372 (37.3%) | 48 (19%) |  |
| 65+ | 67 (5.4%) | 63 (6.3%) | 4 (1.6%) |  |
| Population sector |  |  |  |  |
| Arab | 86 (6.9%) | 57 (5.7%) | 29 (11.5%) | 0.0019 |
| Jewish | 1164 (93.1%) | 941 (94.3%) | 223 (88.5%) |  |
| Socioeconomic status |  |  |  |  |
| Low | 137 (11%) | 94 (9.4%) | 43 (17.1%) | <0.0001 |
| Middle | 448 (35.8%) | 337 (33.8%) | 111 (44%) |  |
| High | 665 (53.2%) | 567 (56.8%) | 98 (38.9%) |  |
| Hospital |  |  |  |  |
| Ha'emek | 160 (12.8%) | 148 (14.8%) | 12 (4.8%) | <0.0001 |
| Kaplan | 133 (10.6%) | 120 (12%) | 13 (5.2%) |  |
| Meir | 218 (17.4%) | 190 (19%) | 28 (11.1%) |  |
| Rabin | 253 (20.2%) | 193 (19.3%) | 60 (23.8%) |  |
| Schneider | 135 (10.8%) | 99 (9.9%) | 36 (14.3%) |  |
| Soroka | 351 (28.1%) | 248 (24.8%) | 103 (40.9%) |  |
| Occupation |  |  |  |  |
| Physician | 234 (18.7%) | 219 (21.9%) | 15 (6%) | <0.0001 |
| Nurse | 549 (43.9%) | 420 (42.1%) | 129 (51.2%) |  |
| Administration and support staff | 467 (37.4%) | 359 (36%) | 108 (42.9%) |  |
| **Frequency of contact with suspected or confirmed Covid-19 patients** |  |  |  |  |
| Always | 63 (5%) | 49 (4.9%) | 14 (5.6%) | 0.0735 |
| Often | 124 (9.9%) | 98 (9.8%) | 26 (10.3%) |  |
| Sometimes | 232 (18.6%) | 196 (19.6%) | 36 (14.3%) |  |
| Seldom | 328 (26.2%) | 259 (26%) | 69 (27.4%) |  |
| Never | 488 (39%) | 388 (38.9%) | 100 (39.7%) |  |
| Unknown | 15 (1.2%) | 8 (0.8%) | 7 (2.8%) |  |
| **Clinical worker with direct patient contact** |  |  |  |  |
| Yes | 725 (58%) | 580 (58.1%) | 145 (57.5%) | 0.3360 |
| No | 492 (39.4%) | 395 (39.6%) | 97 (38.5%) |  |
| Unknown | 33 (2.6%) | 23 (2.3%) | 10 (4%) |  |
| **No. of risk factors according to CDC criteria** |  |  |  |  |
| 0 | 445 (35.6%) | 347 (34.8%) | 98 (38.9%) | 0.0151 |
| 1 | 473 (37.8%) | 374 (37.5%) | 99 (39.3%) |  |
| 2 | 212 (17%) | 167 (16.7%) | 45 (17.9%) |  |
| 3 | 76 (6.1%) | 68 (6.8%) | 8 (3.2%) |  |
| 4+ | 44 (3.5%) | 42 (4.2%) | 2 (0.8%) |  |
| **No. of influenza vaccinations during previous 5 years** |  |  |  |  |
| 0 | 226 (18.1%) | 110 (11%) | 116 (46%) | <0.0001 |
| 1-2 | 453 (36.2%) | 364 (36.5%) | 89 (35.3%) |  |
| 3-4 | 270 (21.6%) | 242 (24.2%) | 28 (11.1%) |  |
| 5+ | 301 (24.1%) | 282 (28.3%) | 19 (7.5%) |  |
| **CDC "certain" risk criteria** |  |  |  |  |
| 65+ years | 67 (5.4%) | 63 (6.3%) | 4 (1.6%) | 0.0048 |
| Cancer | 34 (2.7%) | 29 (2.9%) | 5 (2%) | 0.5572 |
| Chronic Kidney Disease | 59 (4.7%) | 50 (5%) | 9 (3.6%) | 0.4260 |
| Chronic Obstructive Pulmonary Disease | 6 (0.5%) | 6 (0.6%) | 0 (0%) | 0.4692 |
| Heart Disease | 30 (2.4%) | 28 (2.8%) | 2 (0.8%) | 0.1022 |
| Solid-Organ Transplantation | 0 (0.0%) | 0 (0.0%) | 0 (0.0%) |  |
| Obesity: BMI, 30-40 kg/m<sup>2</sup> | 197 (15.8%) | 161 (16.1%) | 36 (14.3%) | 0.5339 |
| Severe obesity: BMI >40 kg/m<sup>2</sup> | 15 (1.2%) | 9 (0.9%) | 6 (2.4%) | 0.1089 |
| Pregnancy | 26 (2.1%) | 2 (0.2%) | 24 (9.5%) | <0.0001 |
| Sickle Cell Disease | 0 (0.0%) | 0 (0.0%) | 0 (0.0%) |  |
| Smoking | 108 (8.6%) | 85 (8.5%) | 23 (9.1%) | 0.8552 |
| Type 2 Diabetes Mellitus | 64 (5.1%) | 58 (5.8%) | 6 (2.4%) | 0.0406 |
| **CDC "possible" risk criteria** |  |  |  |  |
| Asthma | 77 (6.2%) | 63 (6.3%) | 14 (5.6%) | 0.7642 |
| Cerebrovascular Disease | 18 (1.4%) | 18 (1.8%) | 0 (0%) | 0.0641 |
| Other Respiratory Disease | 1 (0.1%) | 1 (0.1%) | 0 (0%) | 0.9999 |
| Hypertension | 111 (8.9%) | 105 (10.5%) | 6 (2.4%) | 0.0001 |
| Immunosuppression | 37 (3%) | 34 (3.4%) | 3 (1.2%) | 0.0996 |
| Neurologic Disease | 51 (4.1%) | 43 (4.3%) | 8 (3.2%) | 0.5255 |
| Liver Disease | 25 (2%) | 20 (2%) | 5 (2%) | 0.9999 |
| Overweight: BMI, 25-30 kg/m<sup>2</sup> | 380 (30.4%) | 313 (31.4%) | 67 (26.6%) | 0.1627 |
| Thalassemia | 10 (0.8%) | 9 (0.9%) | 1 (0.4%) | 0.6830 |
| Type 1 Diabetes mellitus | 5 (0.4%) | 3 (0.3%) | 2 (0.8%) | 0.5826 |

Abbreviations: IQR, interquartile range; CDC, Centers for Disease Control and Prevention; BMI, body mass index; kg, kilograms; m<sup>2</sup>, meters squared.

For the primary analysis, fully vaccinated individuals contributed 68,574 PDs and unvaccinated participants contributed 10,027 PDs. Vaccinated individuals were followed up for a median of 71 days (IQR: 67-76), while unvaccinated participants were followed up for a median of 35 days (IQR: 15-52). Vaccinated participants provided weekly respiratory specimens for 85.9% (standard deviation [SD]: 19.9) of the weeks of follow-up time, while unvaccinated participants contributed respiratory specimens for 88.9% (SD: 17.4) of the follow-up weeks.

There were 13 PCR-positive events during the follow-up period for the primary analysis, including 4 among vaccinated participants (incidence: 0.58 per 10,000 PDs) and 9 among unvaccinated participants (incidence: 8.98 per 10,000 PDs). The adjusted VE was 94.5% (95% CI: 82.6%-98.2%). Table 2 describes VE estimates, using calendar time as the model time scale, for the primary analyses and the secondary analyses; Supplementary Table 3 describes the same estimates using the time-varying analysis.

**Table 2.** Covid-19 vaccine effectiveness for primary and secondary analyses, with calendar time as the model time-scale*

| Analysis | Vaccinated |  |  | Unvaccinated |  |  | VE |  |
| --- | --- | --- | --- | --- | --- | --- | --- | --- |
|  | Person-days | Positive cases | Incidence rate per 10,000 person-days | Person-days | Positive cases | Incidence rate per 10,000 person-days | Unadjusted (1-HR) | Adjusted (1-HR) |
| **Two-dose VE against any infection** | 68,574 | 4 | 0.58 | 10,027 | 9 | 8.98 | 95.4% (84.8%-98.6%) | 94.5% (82.6%-98.2%) |
| **Two-dose VE against symptomatic infection** | 68,605 | 2 | 0.29 | 10,027 | 9 | 8.98 | 97.4% (88%-99.4%) | 97% (72%-99.7%) |
| **Two-dose VE against any infection (14 days after second dose)** | 61,620 | 4 | 0.65 | 10,027 | 9 | 8.98 | 95.4% (84.8%-98.6%) | 94.5% (82.5%-98.2%) |
| **Two-dose VE against any infection including participants regardless of how many weekly specimens they provided (Extended Cohort 1)\*\*** | 67,230 | 4 | 0.59 | 9,229 | 9 | 9.75 | 95.6% (85.6%-98.7%) | 94.8% (83.4%-98.4%) |
| **Two-dose VE against any infection, excluding only participants who were seropositive at enrollment and/or PCR-positive before or at enrollment (Extended Cohort 2)** | 75,388 | 6 | 0.80 | 10,888 | 10 | 9.18 | 94% (83.3%-97.8%) | 92.3% (80.5%-96.9%) |

Notes:
\*All VE analyses were performed for two-dose vaccination with follow-up beginning 7 days after second vaccine unless specified otherwise.
\*\*Extended Cohort 1 included participants who had a negative enrollment serology and were not previously infected by PCR, regardless of how many weekly specimens they provided during the follow-up period.
Abbreviations: VE, vaccine effectiveness; HR, hazards ratio; PCR, polymerase chain reaction

Among the 13 PCR-positive events, 11 were symptomatic and 2 were asymptomatic; none of the symptomatic participants required hospitalization. Both asymptomatic infections were among vaccinated participants. Adjusted VE for symptomatic infection was 97.0% (95% CI: 72.0%-99.7%). VE against asymptomatic infection could not be estimated due to the low number of events during the follow-up period.

In the VE analysis that used a combined outcome of PCR and/or seroconversion, 983 vaccinated participants and 35 unvaccinated participants were included, and contributed 50,526 PDs and 1542 PDs, respectively. During the two-month follow-up period, there were eight infections, of which five were among vaccinated participants (Supplementary Table 4). Unadjusted VE for the combined endpoint was 94.5% (95% CI: 63.0%-99.0%).

Results of secondary analyses largely mirrored the results of the primary VE analysis. VE against any infection among fully vaccinated participants who were at least 14 days after their second dose was identical to the primary analysis for vaccination after 7 days, because all 4 cases among vaccinated participants occurred more than 14 days after their second dose. Adjusted two-dose VE for the Extended Cohort 1 was similar (94.8% [95% CI: 83.4%-98.3%]). Adjusted two-dose VE for Extended Cohort 2 was 89.5% (95% CI: 72.8%-95.9%).

Three samples from infections identified in the primary analysis, and two samples from infections identified among vaccinated participants in the period between the first and the second dose, underwent genetic sequencing and were determined to be alpha variant (B.1.1.7).

## Discussion

We found that two doses of Pfizer BNT162b2 mRNA vaccine were 94.5% effective in preventing a combined endpoint of symptomatic and asymptomatic PCR-confirmed SARS-CoV-2 infection in an infection-naïve cohort of hospital-based HCP in Israel. When we added SARS-CoV-2 seroconversion as an outcome in a secondary analysis, VE remained equally high, at 94.5%. Our findings provide further evidence to results from previous studies in Israel,^18^ the UK^19^ and the US^20^ that have demonstrated the high effectiveness of two doses of the Pfizer BNT162b2 mRNA vaccine in preventing infection among HCP and frontline workers. While our study population was smaller than similar prospective VE studies among HCP and essential workers in the UK and the US, respectively,^6,19,21^ we collected respiratory samples weekly, with high compliance among both vaccinated and unvaccinated participants. Additionally, we collected baseline, one-month and 3-month serology, which we used to both to exclude individuals who had previous infection and to identify individuals who were infected during the study period. Neither the UK nor the US studies used serology to identify new infections, and while the UK study conducted twice weekly asymptomatic testing of frontline HCP using a lateral flow device, routine PCR testing of all participants was conducted once every two weeks.

Asymptomatic infection has been shown to account for nearly half of all SARS-CoV-2 infections^22^ and is likely an important driver of virus transmission in the current pandemic.^23^ However, in our study, despite rigorous weekly collection of respiratory specimens from all participants during the three-month follow-up period, with over 85% compliance, we identified only 2 PCR-positive asymptomatic cases, and therefore we were not able to perform a stratified analysis to evaluate PCR-confirmed asymptomatic infection. The relatively low percentage of asymptomatic cases among all cases [2/13 (15%)] may reflect the fact that we followed up on participants’ symptoms more thoroughly than in other studies,^22^ in which symptom screening was often administered only at the time of testing; in addition to our weekly symptom questionnaire, we contacted all PCR-positive participants after their confirmed infection to ask them whether they had symptoms before or after their positive test. Viral shedding can be as short as a few days in asymptomatically infected individuals,^7^ and therefore our weekly sampling scheme may have missed transient asymptomatic infections. However, by including seroconversion in our secondary analysis, we were likely to identify any cases that were missed by weekly PCR testing. The fact that only one participant without a positive RT-PCR test seroconverted during the study period, and the VE for the joint serology/PCR endpoint was 94.5%, is further reassuring about the effectiveness of two doses of vaccine against asymptomatic infection.

In our study, the five PCR-confirmed SARS-CoV-2 cases that were sequenced were alpha variant, and during the study period, the alpha variant predominated in the country. In Israel, during the study period, the NVL sequenced 11,452 samples, of which 8,116 (70.9%) were alpha variant; 2552 (22.3%) were wild-type virus; the remainder were a mix of variants, including beta (268[2.3%]) and those from the B.1.617 family [39(0.3%)] (personal communication, Michal Mandelboim and Neta Zuckerman, Israel NVL). Our study, therefore, largely addresses VE against the alpha variant, and adds more evidence to previous studies that have demonstrated high two-dose VE for the Pfizer BNT162b2 mRNA against the alpha variant in the initial months following vaccination.^15,24,25^

Strengths of our study include the weekly collection of respiratory specimens, use of serology testing to identify both previous infection and new infections, and the use of the CHS EMR characterize participants’ demographic and clinical history, Covid-19 and influenza vaccine history, and to identify prior and new SARS-CoV-2 infections.

Our study has a number of limitations. First, because participation was voluntary, it may suffer from selection bias, which may limit its generalizability; participants who chose to participate likely differed from the broader HCP population and the overall Israeli population in quantifiable and unquantifiable ways. Second, vaccinated and unvaccinated participants differed with respect to a number of demographic characteristics. We did, however, adjust for many of these differences, such as sex, SES and occupation in our analyses. Third, we used different combinations of serology testing to determine enrollment serological status. Differences in sensitivities of different test combinations may have created inconsistencies in serological status determination. In addition, while the serological test (Abbott SARS-CoV-2 nucleocapsid IgG test) we used to determine new infections during the 30- to 90-day follow-up window has been shown to be over 90% sensitive in identifying PCR infections in mildly symptomatic individuals,^26^ it may be less sensitive in capturing asymptomatic individuals, and if so we may have missed some asymptomatic cases.

In conclusion, in our prospective study of HCP across six hospitals in Israel with rigorous weekly surveillance, we found very high VE following two doses of Pfizer BNT162b2 mRNA vaccine against both symptomatic and asymptomatic SARS-CoV-2 during a period of predominant alpha variant circulation. While these results are encouraging, continued monitoring of Covid-19 VE in Israel and globally is critical to monitor the duration of VE, to evaluate VE against emerging variants of concern, and to inform decision-making about the need for booster doses.

## Data Availability

Because of data privacy regulations, the raw data for this study cannot be publicly shared.

## Trial Registration

ClinicalTrials.gov Identifier: NCT04709003

## Author Contributions

MAK., EBH., BC, MC, DG, AP, ST, JL, MY, AH., EK, ND, NB. and RDB conceived the study and developed the study methods.

EBH, BC, MC, DG, AP, ST, JL, MY, AH and DR collected the data.

DA, RW, ABD, EKT, CBC, DS, HBT, RB, GR, YSA, MM, NZ and NR analyzed the laboratory data.

JL, EK, AA, ND, NB, MAK, and MY conducted the analysis

JL and EK verified the data from the CHS EMR, and EBH, BC, MC, DG, AP, ST, JL, and MY verified the data collected at the study sites.

MAK, JL, MY, AH, ND, NB, EK and RDB wrote the first draft of the manuscript All authors revised the manuscript and edited the final manuscript.

## Data Sharing Statements

Because of data privacy regulations, the raw data for this study cannot be publicly shared.

## Acknowledgements

We thank Nadav Raviv and Ilan Gofer from Clalit Research Institute, Innovation Division for helping with the data collection throughout the study, Sydney Krispin from Clalit Research Institute, Innovation Division for helping with the manuscript submission. We also thank Arnold Monto, Emily Martin, Josh Petrie and Ryan Malosh from University of Michigan School of Public Health for providing input on the analysis plan. We thank Yossef Rosemberg from Clalit Central laboratory for his help with the serology testing. We thank Svetlana Rothe, Ranin Wahab, Tamar Amar and Tal Eylon from Rabin Medical Center, Anna Yanovskay, Carmel Kasher, Merav Strauss and Hana Kahanov Edelstein from Ha’Emek Medical Center, Ayman Fadeela and Galit Carmon from Meir Medical Center, Iris Greenbaum, Nataliya Kyrylyshyn, Yosfit Yosefa Mussa, Abeer Bdeer, Inbar Gesua Abu from Schneider Children’s Medical Center of Israel, Ayelet Sherman, Shlomit shick, Lihi Geler and Hana Leiba from Kaplan Medical Center, Orli Zamir- Barashi and the pediatric infectious disease unit from Soroka University Medical Center for recruiting participants, collecting samples and following-up on participant questionnaires during the study. We thank the Israeli Ministry of Health for providing serology kits for the 30-day and 90-day serology tests. We also thank all the participants for their contribution throughout the study.

## Supplementary Appendix

### Supplementary Methods 1. Description of Genetic Sequencing Performed at Israel National Virology Laboratory and Samir Medical Center

The Israel NVL performed whole genome sequencing using the COVID-seq library preparation kit (Illumina). Library validation and mean fragment size was determined by TapeStation 4200 via DNA HS D1000 kit (Agilent). Libraries were pooled, denatured and diluted to 10pM and sequenced on NovaSeq (Illumina). Resulting fastq files were subjected to quality control using FastQC (Babraham Bioformatics, UK) and MultiQC^27^ and low-quality sequences were filtered using trimmomatic.^28^ Sequences were mapped to the SARS-CoV-2 reference genomes (NC_045512.2) using BWA mem.^29^ Resulting BAM files were sorted and indexed using the SAMtools suite.^30^ Consensus fasta sequences were assembled using iVar^31^, with positions with <5 nucleotides determined as Ns. Multiple alignment of sample sequences with SARS-CoV-2 reference genome (NC_045512.2) was done with MAFFT.^32^ Variant-specific mutations were identified via a custom python script. At AHMC, the methods were identical, except that NextSeq was used instead of Noveseq.

### Supplementary Methods 2. Algorithm for determining serological status for enrollment serology, 30-day serology and 90-day serology, CoVEHPI

The cutoff values described below for positive, negative and indeterminant for the three tests were determined according to the manufacturers’ package insert.

1. Definitions of enrollment serology results (positive, negative, indeterminant)
  a. Positive
    i. Abbott Architect N ≥ 1.4 regardless of the Diasorin or the Quant test
    ii. For unvaccinated participants, the following will also be considered positive:
      1. Abbott SARS-CoV-2 IgG II Quant ≥ 50.0 regardless of results for Abbott Architect N
      2. Diasorin ≥ 15.0 regardless of results for Abbott Architect N
  b. Negative
    i. Abbott Architect N < 0.49 AND Diasorin < 12.0
    ii. Abbott Architect N < 0.49 without a Diasorin test or a Quant test
    iii. Abbott SARS-CoV-2 IgG II Quant <50 without another test
    iv. Diasorin < 12.0 without an Abbott Architect N test
    v. Abbott Architect N < 0.49 AND Quant test < 50
    vi. For vaccinated participants >7 days after first dose
      1. Abbott SARS-CoV-2 IgG II Quant ≥ 50.0 AND Abbott Architect N < 0.49
      2. Diasorin ≥ 12.0 positive AND Abbott Architect N <0.49
  c. Indeterminant in
    i. unvaccinated participant or participant ≤ 7 days after first vaccine
      1. Architect N 0.49--1.39 without Diasorin test OR without Abbott SARS-CoV-2 IgG II Quant
      2. Architect N 0.49--1.39 with Diasorin test < 12.0 OR with Abbott SARS-CoV-2 IgG II Quant <50 (Negative)
      3. Diasorin 12.0-15 without an Abbott Architect N test (only for unvaccinated participants or participants <7 days after first dose)
      4. Diasorin 12.0-15 with an Abbott Architect N test <1.40
    ii. vaccinated person > 7 days after first dose
      1. Architect N 0.49--1.39 with Diasorin test ≥ 12.0 OR with Abbott SARS-CoV-2 IgG II Quant ≥50
      2. Architect N (0.49--1.39 without Diasorin test OR Abbott SARS-CoV-2 IgG II Quant
      3. Architect N 0.49--1.39 with Diasorin test < 12 OR Abbott SARS-CoV-2 IgG II Quant <50 (negative)
  d. Irrelevant serology results –Vaccinated participants who had serology collected > 7 days after first vaccine will be excluded from the primary analysis if they only have the following tests :
    i. Abbott SARS-CoV-2 IgG II Quant ≥ 50.0 without results for Abbott Architect N
    ii. Diasorin ≥ 12.0 without results for Abbott Architect N
2. Determination of new infections by serology in 30-day to 90-day follow-up period
  a. Definition of New infection by 90-day serology
    i. A participant who is negative by serology at 30 days and positive by serology at 90 days and
  b. Determination of 90-day serology results
    i. Positive. Abbott Architect N ≥ 1.4 regardless of the result of the Quant test AND regardless of the result of Diasorin test
    ii. Negative. Abbott Architect N < 0.49 regardless of the result of the Quant test AND regardless of the result of Diasorin test
    iii. Architect N 0.49--1.39

### Supplementary Methods 3. Sample Size Considerations

In order to estimate the required sample size, we assumed an incidence of SARS-CoV-2 of 5% in unvaccinated participants over a 12-month period, with 80% vaccine coverage. Based on these assumptions, the sample size required to detect a vaccine effectiveness (VE) of 50%, using the Cox Proportional Hazards (PH) model, in order to reach 80% power level with alpha = 0.05, was 3153 participants.

Several weeks into our study, after high post-introduction VE estimates emerged^15,24^, and in light of high transmission rates in Israel,^17^ we reassessed these estimations; we still estimated a vaccine coverage of 80%, but we assumed a three-month attack rate of 3.5% unvaccinated participants, and set the power to detect VE of 85%. Based on these assumptions, 80% power and alpha = 0.05, we revised target sample size to 1300 subjects.

### Supplementary Methods 4. Additional Assessments of Vaccine Effectiveness

For the primary analysis of VE against any infection, and the secondary analyses of VE against symptomatic and asymptomatic infection, we also excluded participants who had not provided a weekly swab for the first 21 days of the follow-up period, because we thought we could not reliably rule out infections in these participants; participants who did not provide weekly swabs for at least 21 days during follow-up were right-censored at the first day of this period.

We also performed sensitivity analyses where we included 1) participants who had not provided a weekly swab for a period of at least 21 days (“Extended Cohort 1”); and 2) participants who had indeterminant enrollment serology results, who had not provided an enrollment serology, or whose serology sample was not tested with a reliable test to exclude previous infection, regardless of how many weekly swabs they had provided (“Extended Cohort 2”).

**Supplementary Table 1.** RT-PCR methods used by hospital laboratories and Clalit Health Services (CHS) central laboratory for testing of enrollment and weekly samples, CoVEHPI

| **Laboratory name** | **Methods** |
| --- | --- |
| **CHS central laboratory, Atidim, Tel Aviv, Israel** | RNA extraction was performed by STAR Hamilton/ I7 Beckman instrument. Real- Time PC R was done using Quant Studio 5 (Thermo qPCR kit) / CFX (Allplex nCOVID-19 kit). Reruns for borderline results were conducted on COBAS 6800 (using the sample-to-result method)<br>In addition, some of the weekly specimens were processed by pooling (2:1) before RNA extraction according to the algorithm from POOLD Diagnostics (Beer-Sheva, Israel); we estimated that these samples met the criteria for pooling because they had a low probability for positive results (less than 6%). |
| **Rabin Medical Center, Petah Tikva, Israel** | RNA extraction and PCR setup was done by STAR/STAR LET Hamilton and by NeuMoDx 288 (sample to result method). Real Time PCR was done by CFX (Allplex nCOVID-19 kit). |
| **Meir Medical Center, Kfar Saba, Israel** | RNA extraction was done by STAR Hamilton (Seegene Starmag Viral RNA kit). Real Time PCR was done by CFX 96 (Seegene Allplex nCOVID-19 kit). Reruns for borderline results were performed on GeneXpert (from sample to result method). |
| **Kaplan Medical Center, Rehovot, Israel** | RNA extraction was done by NIMBUS / STAR Hamilton Real Time PCR was done by CFX (Allplex nCOVID-19 kit/ GENXPRT) |
| **Soroka University Medical Center, Beer Sheva, Israel** | RNA extraction and Covid-19 detection was done using Seegene STARMag extraction kit and SARS-CoV-2 assay amplification kit. Samples from healthy |
|  | population with low probability for positive results (less than 6%) were processed by pooling (2:1) before RNA extraction according to the algorithm used by POOLD Diagnostics (Beersheva, Israel) . |
| **Ha'Emek Medical Center, Afula, Israel** | Real Time PCR was done by Quant Studio 5 (Thermo qPCR kit) / CFX (Allplex nCOVID-19 kit). Reruns for borderline results were done on COBAS 6800 or Hologic Panther (from sample to result method). |

**Supplementary Table 2.** Demographic and clinical characteristics of all enrolled participants (N=1567)

| Characteristic | N (%) |
| --- | --- |
| **Sex, no. (%)** |  |
| Female | 1238 (79%) |
| Male | 329 (21%) |
| Age Median (IQR) | 45 (36-54) |
| **Age group, no. (%)** |  |
| 18-34 | 342 (21.8%) |
| 35-49 | 636 (40.6%) |
| 50-64 | 506 (32.3%) |
| 65+ | 83 (5.3%) |
| **Population sector, no. (%)** |  |
| Arab | 119 (7.6%) |
| Jewish | 1448 (92.4%) |
| **Socioeconomic status, no. (%)** |  |
| Low | 185 (11.8%) |
| Middle | 550 (35.1%) |
| High | 830 (53%) |
| Unknown | 2 (0.1%) |
| **Hospital** |  |
| Haemek | 178 (11.4%) |
| Kaplan | 172 (11%) |
| Meir | 257 (16.4%) |
| Rabin | 301 (19.2%) |
| Schneider | 181 (11.6%) |
| Soroka | 478 (30.5%) |
| **Occupation** |  |
| Physician | 294 (18.8%) |
| Nurse | 694 (44.3%) |
| Administration and support staff | 579 (36.9%) |

| **Frequency of contact with suspected or confirmed Covid-19 patients** |  |
| --- | --- |
| Always | 82 (5.2%) |
| Often | 157 (10%) |
| Sometimes | 280 (17.9%) |
| Seldom | 415 (26.5%) |
| Never | 601 (38.4%) |
| Unknown | 32 (2%) |
| **Clinical worker with direct patient contact** |  |
| Yes | 907 (57.9%) |
| No | 605 (38.6%) |
| Unknown | 55 (3.5%) |
| **No. of risk factors according to CDC criteria** |  |
| 0 | 568 (36.2%) |
| 1 | 578 (36.9%) |
| 2 | 270 (17.2%) |
| 3 | 93 (5.9%) |
| 4+ | 58 (3.7%) |
| **No. of influenza vaccinations during previous 5 years** |  |
| 0 | 294 (18.8%) |
| 1-2 | 560 (35.7%) |
| 3-4 | 344 (22%) |
| 5+ | 369 (23.5%) |
| **CDC "certain" risk criteria, no. (%)** |  |
| 65+ years | 83 (5.3%) |
| Cancer | 38 (2.4%) |
| Chronic Kidney Disease | 88 (5.6%) |
| Chronic Obstructive Pulmonary Disease | 8 (0.5%) |
| Heart Disease | 38 (2.4%) |
| Solid-Organ Transplantation | 0 (0.0%) |
| Obesity: BMI, 30-40 kg/m<sup>2</sup> | 247 (15.8%) |
| Severe obesity: BMI >40 kg/m<sup>2</sup> | 17 (1.1%) |
| Pregnancy | 33 (2.1%) |
| Sickle Cell Disease | 0 (0.0%) |
| Smoking | 136 (8.7%) |
| Type 2 Diabetes Mellitus | 82 (5.2%) |
| **CDC "possible" risk criteria, no. (%)** |  |
| Asthma | 91 (5.8%) |
| Cerebrovascular Disease | 20 (1.3%) |
| Other Respiratory Disease | 1 (0.1%) |
| Hypertension | 141 (9%) |
| Immunosuppression | 52 (3.3%) |
| Neurologic Disease | 56 (3.6%) |
| Liver Disease | 32 (2%) |
| Overweight: BMI, 25 – 30 kg/m<sup>2</sup> | 471 (30.1%) |
| Thalassemia | 11 (0.7%) |
| Type 1 Diabetes mellitus | 8 (0.5%) |
| **Vaccinated with one dose prior to or at enrollment** | **1208 (77.1%)** |
| **Unvaccinated at enrollment** | **359 (22.9%)** |

Abbreviations: IQR, interquartile range; CDC, Centers for Disease Control and Prevention; BMI, body mass index; kg, kilograms; m<sup>2</sup>, meters squared.

**Supplementary Table 3.** Covid-19 Vaccine Effectiveness for primary and secondary analyses, using time-varying analysis*

| Analysis | Vaccinated |  |  | Unvaccinated |  |  | VE |  |
| --- | --- | --- | --- | --- | --- | --- | --- | --- |
|  | Person-days | Positive cases | Incidence rate per 10,000 person-days | Person-days | Positive cases | Incidence rate per 10,000 person-days | Unadjusted (1-HR) | Adjusted (1-HR) |
| **Two-dose VE against any infection** | 68,574 | 4 | 0.58 | 10,027 | 9 | 8.98 | 94% (79.9%-98.2%) | 93.1% (74.1%-98.2%) |
| **Two-dose VE against symptomatic infection** | 68,605 | 2 | 0.29 | 10,027 | 9 | 8.98 | 96.6% (83.9%-99.3%) | 96.2% (50.4%-99.7%) |
| **Two-dose VE against any infection - 14 days after second dose** | 61,620 | 4 | 0.65 | 10,027 | 9 | 8.98 | 93.3% (77.6%-98%) | 91.9% (69.6%-97.9%) |
| **Two-dose VE against any infection including participants regardless of how many weekly specimens they provided \*\*** | 67,230 | 4 | 0.59 | 9,229 | 9 | 9.75 | 94.4% (81.1%-98.3%) | 93.5% (76.3%-98.2%) |
| **Two-dose VE against any infection, excluding only participants who were seropositive at enrollment and/or PCR-positive before or at enrollment** | 75,388 | 6 | 0.80 | 10,888 | 10 | 9.18 | 91.7% (76.6%-97.1%) | 89.5% (72.8%-95.9%) |

Notes:
\*All VE analyses were performed for two-dose vaccination with follow-up beginning 7 days after second vaccine unless specified otherwise.
\*\*Extended Cohort 1 included participants who had a negative enrollment serology and were not previously infected by PCR, regardless of how many weekly specimens they provided during the follow-up period.
Abbreviations: HR, hazards ratio; VE, vaccine effectiveness, PCR, polymerase chain reaction

**Supplementary Table 4.** Concordance between PCR results and serology results in secondary vaccine effectiveness analysis using positive PCR result and/or seroconversion as a combined outcome

| **Vaccination status** | **PCR results** | **Seropositive** | **Indeterminant serology** | **Seronegative** |
| --- | --- | --- | --- | --- |
| **Vaccinated** | PCR-positive | 1 | 2 | 1 |
|  | PCR-negative | 1 | 0 | 0 |
| **Unvaccinated** | PCR-positive | 1 | 2 | 0 |
|  | PCR-negative | 0 | 0 | 0 |

Abbreviation: PCR, polymerase chain reaction.

